# Effects of Working from Home on Musculoskeletal Pain and Coronaphobia during the COVID-19 Pandemic

**DOI:** 10.1101/2025.09.16.25335923

**Authors:** Luciano de Arruda Castelo, Nelson Carvas Junior, Vinicius Tassoni Civile, Juliana Moreira Costa, Rodolfo Ribeiro de Matos, Virgínia Fernandes Moça Trevisani

## Abstract

**Background:** COVID-19 pandemic impacted the physical and mental health of Brazilian workers, especially those who started acting in home-office. Changes in the workplace, increased physical inactivity and fear of contracting the disease, known as coronophobia, may be related to increased musculoskeletal pain.

**Objective:** To analyze the prevalence of musculoskeletal pain and coronophobia in home-office workers during the COVID-19 pandemic, as well as evaluating the relationship between these variables.

**Method:** A cross-sectional, observational and descriptive study with 736 participants was conducted through online questionnaire. Sociodemographic data, working conditions, pain, besides the evaluation of coronophobia using COVID-19 Phobia scale (C19P-S) and pain with the Nordic questionnaire of osteomuscular symptoms (NMQ) were collected.

**Results:** Most participants were female (78%), with an average age of 32.8 ± 10.7 years. During the pandemic, 71% reported some osteomuscular symptoms, and of these, 64% began to have new paintings of pain in the pandemic, with higher incidence in the lower back and neck. Participants who began to perform their functions in home-office presented a higher incidence of pain and, among them, 53% reported that the furniture was not suitable for homework. Most had little fear of Covid-19, but moderate levels of coronophobia increased the chance of musculoskeletal pain (RP=1,74; IC 95%). There was a significant association between coronophobia and musculoskeletal pain.

**Conclusions:** The pandemic raised the prevalence of musculoskeletal pain, especially in cervical and lumbar regions, and the fear of Covid-19 influenced this relationship.

## INTRODUCTION

The SARS-CoV-2 virus, named COVID-19 by the World Health Organization (WHO), was discovered at the end of December 2019 in Wuhan, China. On March 11, 2020, the WHO declared a pandemic due to the uncontrolled spread of the virus worldwide (1). Since its discovery, COVID-19 has affected more than 119 million people globally, with over 2 million confirmed deaths (2), significantly impacting public health and the economy of several countries. In addition to the direct health consequences, the pandemic also triggered various side effects, including the emergence of new psychological conditions and the worsening of pre-existing ones.

Preventive measures such as social isolation, hand and food hygiene, and mask use were necessary to reduce transmission (3). In Brazil, in March 2020, a quarantine bill was approved (4), which defined the isolation of healthy individuals and those in the incubation period of the disease, i.e., individuals presenting symptoms or testing positive for COVID-19. This physical distancing allowed better control and a reduction in contamination rates (5). At the time, there was no definitive treatment, and vaccines only began to be administered as a control measure in January 2021.

Home isolation was one of the strategies adopted to control COVID-19 transmission, serving as an effective way to prevent exposure to infection and reduce fatality rates (6). This strategy, whether voluntary or mandatory, while effective, could also lead to physical inactivity, weight gain, behavioral dependence, and social isolation (7). Previous studies indicated that prolonged isolation could be associated with psychological disorders (7), sleep disturbances (8), severe phobias (9,10), musculoskeletal conditions and pain (11), the latter also being exacerbated by physical inactivity (12). Due to the social isolation imposed by the pandemic, many individuals were compelled to work from home and adopt a home-office regimen (14).

Studies on the prevalence of musculoskeletal pain during the pandemic are still scarce, particularly among individuals who transitioned to working from home. Similarly, few studies assess the phobias acquired during the pandemic, especially coronavirus-related phobia, known as coronaphobia (10), defined as the excessive and irrational fear of contracting COVID-19. This condition has been widely observed in the general population, leading to significant increases in anxiety, stress, and other psychological disorders (13). Coronaphobia can trigger several physical responses, including increased muscle tension and musculoskeletal pain (11).

The relationship between coronaphobia and musculoskeletal pain can be understood through the impact of chronic stress on the human body (11). The constant fear of contracting COVID-19 and concerns about its consequences may lead to prolonged muscle tension, which in turn contributes to the development of musculoskeletal pain, particularly in areas such as the neck, shoulders, and back (14).

This situation is particularly relevant for workers who adopted a home-office routine during the pandemic. The sudden shift to remote work, often without adequate preparation of the home environment, resulted in ergonomically inadequate conditions for many. Moreover, the combination of emotional stress caused by coronaphobia and the lack of a proper work environment increased the incidence of musculoskeletal pain among this population (15,16).

Therefore, this study aims to analyze the prevalence of musculoskeletal pain and coronaphobia in home-office workers during the COVID-19 pandemic, as well as to evaluate the relationship between these variables. Understanding these factors is essential for developing more effective intervention strategies to mitigate these negative effects and promote workers’ health and well-being during times of public health crisis.

## METHOD

This was a cross-sectional, observational, and descriptive study, approved by the Research Ethics Committee of Universidade Paulista / UNIP (Protocol: 46136621.3.0000.5512). The study was based on data collected through an online questionnaire addressing perceived changes in workers’ routines before the pandemic, when they worked in a usual environment, and after isolation, when most transitioned to working from home.

The questionnaire was created using Google Forms and was available at the link: https://docs.google.com/forms/d/e/1FAIpQLSdTII48kx_tX1owfMVvb870ODJGfW85xWlXsRt7FocgJnk81w/viewform.

It remained open for three months or until a minimum of 700 responses was reached, following the study by Celenay et al. (2020) (11). Data collection lasted 3 months and 24 days.

The questionnaire was self-administered, completed by participants without assistance, and disseminated through social media (Facebook® and Instagram®), messaging apps (WhatsApp®), and email.

The sample consisted of individuals who voluntarily completed the questionnaire after agreeing to data collection and providing informed consent (ICF). Responses considered valid were from employed individuals of both sexes, aged 18 years or older. Incomplete or inconsistent questionnaires were excluded.

The variables studied included age, sex, height, weight, marital status, education, occupation/profession, presence of chronic disease or health risk factor, regular physical activity before and during the pandemic, workplace during the pandemic, presence of pain before and during the pandemic, pain location and characteristics, and features related to coronaphobia.

For musculoskeletal pain, the Nordic Musculoskeletal Questionnaire (NMQ) was used, which assesses symptoms in nine body regions: neck, upper back, lower back, shoulders, elbows, wrists/hands, hips/thighs, knees, and ankles/feet (17).

Coronaphobia was assessed using the COVID-19 Phobia Scale (C19P-S), developed by Arpaci et al. (2021) (13) and adapted for Brazil by Faro et al. (2022) (18). The scale consists of seven questions rated on a 5-point scale ranging from “strongly disagree (1)” to “strongly agree (5).” Higher scores indicate greater coronaphobia and fear of the disease. The total score ranges from 7 to 35 points. For analysis, scores were stratified into three categories of approximately 33.3% each: 7–19 points = “low fear,” 20–26 points = “moderate fear,” and ≥27 points = “high fear” (13).

Statistical analysis was performed using R software version 4.0 for Mac iOS. Descriptive statistics were used to calculate means and standard deviations for quantitative variables, and absolute numbers and percentages for categorical variables. Group comparisons were performed using Mann-Whitney tests, and associations between categories and groups were analyzed using Pearson’s chi-square and Fisher’s exact tests. Prevalence ratios (PR) with 95% confidence intervals (CI) were calculated using Poisson regression models with robust variance, assuming an alpha error of 0.05. Musculoskeletal pain (from the NMQ) was adopted as the dependent variable. Independent variables included demographic characteristics (sex and age), obesity, and physical activity. Poisson regression models began with a parsimonious model (dependent + one independent variable), followed by an adjusted model including participant characteristics (sex and age). Model fit was evaluated using Akaike Information Criterion (AIC), while the significance of associated factors (obesity and physical activity) was determined at p < 0.05.

## RESULTS

A total of 1,053 individuals responded to the questionnaire. However, 317 respondents were excluded (75 for not meeting the age eligibility criteria and 242 for being unemployed at the time of completion) (Figure 1).

**Figure 1.**
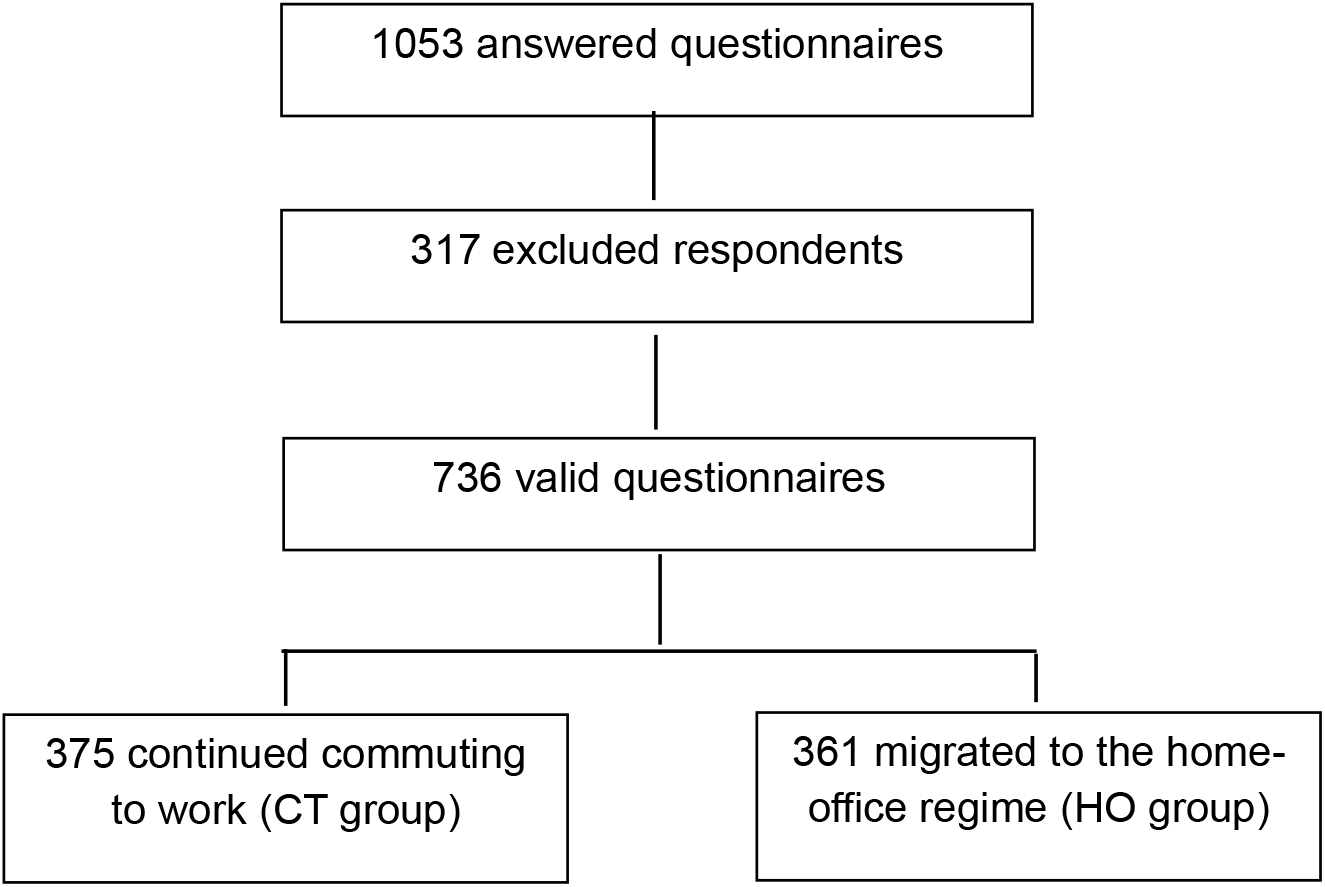
Flow diagram of participant recruitment

The final sample consisted of 736 participants, the majority female (78%), with a mean age of 32.8 ± 10.7 years. More than half were single (57%) and had completed high school. Table 1 presents the sociodemographic characteristics of participants according to their workplace during the pandemic.

**Table 1.** Sociodemographic characteristics of participants in the CT and HO groups.

| Variable | Groups |  |  | p value<sup>2</sup> |
| --- | --- | --- | --- | --- |
|  | CT, N = 375<sup>1</sup> | HO, N = 361<sup>1</sup> | Total, N = 736<sup>1</sup> |  |
| Age, years | 31.5 (10.4) | 34.1 (10.9) | 32.8 (10.7) | 0.001 |
| Weight, kg | 73.4 (16.8) | 72.4 (16.5) | 72.9 (16.7) | 0.393 |
| Height, m | 1.7 (0.1) | 1.7 (0.1) | 1.7 (0.1) | 0.225 |
| BMI, Kg/m<sup>2</sup> | 26.4 (5.2) | 26.3 (5.1) | 26.3 (5.1) | 0.834 |
| Sex, n (%) |  |  |  | 0.283 |
| Female | 288 (77%) | 289 (80%) | 577 (78%) |  |
| Male | 87 (23%) | 72 (20%) | 159 (22%) |  |
| Marital status |  |  |  | 0.038 |
| Married | 125 (33%) | 146 (40%) | 271 (37%) |  |
| Divorced | 18 (4.8%) | 27 (7.5%) | 45 (6.1%) |  |
| Single | 230 (61%) | 186 (52%) | 416 (57%) |  |
| Widower | 2 (0.5%) | 2 (0.6%) | 4 (0.5%) |  |
| Education |  |  |  | <0.001 |
| Elementary school | 6 (1.9%) | 1 (0.3%) | 7 (1.1%) |  |
| High school | 143 (45%) | 103 (32%) | 246 (38%) |  |
| Bachelor's Degree | 86 (27%) | 95 (30%) | 181 (28%) |  |
| Postgraduate | 86 (27%) | 122 (38%) | 208 (32%) |  |
| Comorbidity |  |  |  | 0.136 |
| No | 290 (77%) | 262 (73%) | 552 (75%) |  |
| Yes | 85 (23%) | 99 (27%) | 184 (25%) |  |

<sup>1</sup> Mean (SD); N (%)
<sup>2</sup> Wilcoxon rank sum teste; Pearson's Chi-squared test; Fisher's exact test

Overall, participants who transitioned to home-office (HO group) were significantly older (p = 0.001) than those who continued commuting to work (CT group). The groups also differed in marital status (p = 0.038) and educational level (p < 0.001).

During the pandemic, 71% of participants reported musculoskeletal symptoms, and among these, 64% developed new pain conditions during this period. Participants in the HO group showed a higher incidence of pain compared to the CT group (Table 2).

**Table 2.** Pain before and during the COVID-19 lockdown in the CT and HO groups.

| Variable | Groups |  |  | p value<sup>2</sup> |
| --- | --- | --- | --- | --- |
|  | CT, N = 375<sup>1</sup> | HO, N = 361<sup>1</sup> | Total, N = 736<sup>1</sup> |  |
| Pain before the pandemic |  |  |  | 0.015 |
| No | 143 (59%) | 192 (69%) | 335 (64%) |  |
| Yes | 101 (41%) | 87 (31%) | 188 (36%) |  |
| Pain during the pandemic |  |  |  | <0.001 |
| No | 131 (35%) | 82 (23%) | 213 (29%) |  |
| Yes | 244 (65%) | 279 (77%) | 523 (71%) |  |

<sup>1</sup> N (%)
<sup>2</sup> Pearson's Chi-squared test

The most frequently affected regions during lockdown were the lower back and neck, while the least affected region was the elbows (Figure 2). HO participants reported more elbow pain (17.9% vs. 10.7%, p = 0.019) and wrist pain (35.5% vs. 27.0%, p = 0.038) than CT participants. Conversely, CT participants reported more ankle pain (26.6% vs. 15.1%, p = 0.001) than HO participants (Figure 2).

**Figure 2.**
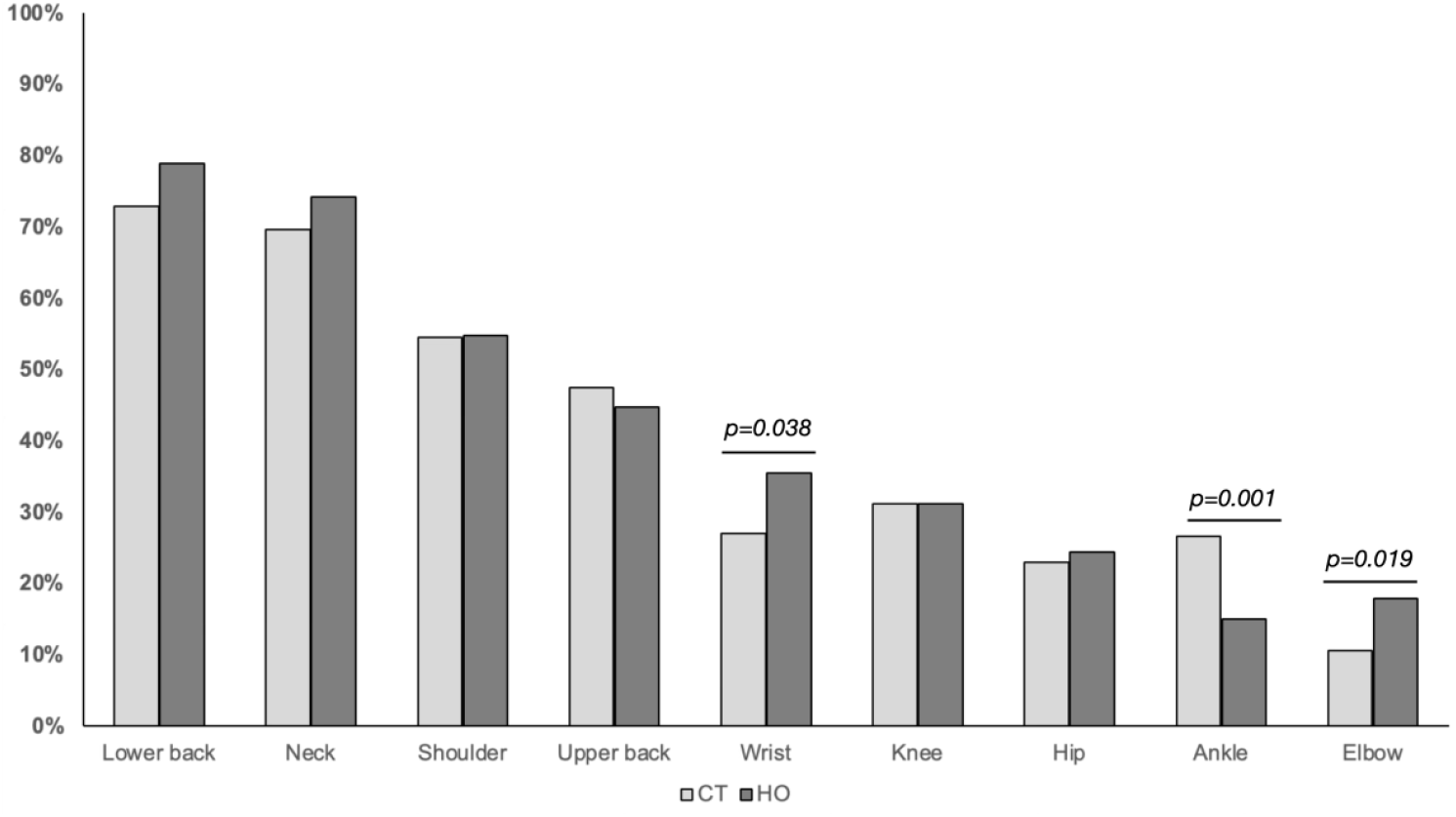
Pain frequency in CT and HO groups by assessed body region

Among the 361 HO participants, 53% reported inadequate home furniture for work. However, no statistically significant differences were found in the frequency of pain by body region between those who considered their furniture adequate versus inadequate (Table 3).

**Table 3.** Comparison of musculoskeletal pain location according to perceived furniture suitability (adequate vs. inadequate)

| Body region | Perception of work furniture |  | p value<sup>2</sup> |
| --- | --- | --- | --- |
|  | Inadequate, N = 163<sup>1</sup> | Adequate, N = 116<sup>1</sup> |  |
| Neck | 124 (76%) | 83 (72%) | 0.395 |
| Shoulder | 89 (55%) | 64 (55%) | 0.925 |
| Upper back | 75 (46%) | 50 (43%) | 0.630 |
| Elbow | 25 (15%) | 25 (22%) | 0.182 |
| Lower back | 131 (80%) | 89 (77%) | 0.463 |
| Wrist | 61 (37%) | 38 (33%) | 0.422 |
| Knee | 55 (34%) | 32 (28%) | 0.274 |
| Hip | 42 (26%) | 26 (22%) | 0.520 |
| Ankle | 24 (15%) | 18 (16%) | 0.855 |

<sup>1</sup> N (%)
<sup>2</sup> Pearson's Chi-squared test

Regarding COVID-19 Phobia results, most participants reported “low fear” (64%), followed by “moderate fear” (25%) and “high fear” (11%) (Table 4).

**Table 4.** COVID-19 Phobia Scale vs. pain occurrence during the pandemic.

|  | YES<br>523 (71%) | NO<br>213 (29%) | TOTAL<br>736 (100%) | p value<sup>1</sup> |
| --- | --- | --- | --- | --- |
| **COVID-19 Phobia Scale** |  |  |  | 0.023 |
| Low fear (*7 to 19 points*) | 318 (61%) | 151 (71%) | 469 (64%) |  |
| Moderate fear (*20 to 26 points*) | 143 (27%) | 39 (18%) | 182 (25%) |  |
| High Fear (*above 27 points*) | 62 (12%) | 23 (11%) | 85 (11%) |  |

<sup>1</sup>Pearson ´s Chi-squared test

Participants with moderate fear were 74% more likely to experience pain during the pandemic compared to those with low fear (Table 5).

**Table 5.** Association between coronaphobia and musculoskeletal pain during the pandemic.

|  | n / N | Prevalence | PR<br>(CI 95%) | p value<sup>1</sup> |
| --- | --- | --- | --- | --- |
| **COVID-19 Phobia Scale** |  |  |  |  |
| Low fear (*7 to 19 points*) | 318 / 523 | 61% | 1,00 |  |
| Moderate fear (*20 to 26 points*) | 143 / 523 | 27% | 1,74 (1,17 - 2,63) | 0,007 |
| High Fear (*above 27 points*) | 62 / 523 | 12% | 1,28 (0,77 - 2,18) | 0,348 |

<sup>1</sup> Poisson regression

## DISCUSSION

During the COVID-19 pandemic, several measures were implemented to reduce transmission, including social and physical isolation. This strategy, although effective, led to physical inactivity, weight gain, behavioral changes, and musculoskeletal conditions. One of the main drivers of these changes was the compulsory shift to home-office (14).

A study published in 2021 reported an increase in physical health complaints, with approximately 64.8% of respondents stating they had developed some health-related issue when working from home (19). Home-office often involves sedentary computer work in non-ergonomic environments, which can contribute to health problems, including musculoskeletal pain (MSP) (11). Bosma et al. (2023) (15) concluded that remote workers were at greater risk of developing MSP during the first year of the COVID-19 pandemic.

The present study found a high prevalence of MSP and an increase in new pain cases among Brazilian home-office workers during the pandemic, reinforcing previous findings (14,15,16,20). Statistical analyses also revealed significant associations between sociodemographic variables, working conditions, and mental health.

The sample, composed of 736 individuals, predominantly female, with a mean age of 32.8 years, represents a typical profile of young, working-age adults. According to Vargová et al. (2023) (21), pandemic-related anxiety manifests differently across age groups, being more reactive and intense among young adults, who often face greater uncertainty regarding financial, academic, and professional futures. The older age of participants in the HO group suggests that this group may have had greater ability or necessity to adapt to remote work, possibly due to professional experience or family responsibilities. Differences in marital status and educational level also suggest sociodemographic influences on adaptation to remote work and potential MSP incidence.

Regarding MSP, the occurrence of new episodes was significantly associated with the pandemic, pain worsening compared to the pre-pandemic period, and home-office. The high prevalence of musculoskeletal symptoms underscores the impact of changes in the work environment on physical health. These findings are consistent with Loef et al. (2022) (22), who identified sedentary behavior as a mediator between remote work and MSP. Ergonomics and postural habits, therefore, become key factors in pain development among remote workers, as also reported by Celenay et al. (2020) (11). The fact that more than half of participants reported inadequate furniture further suggests that domestic environments were not properly prepared for long-term work, contributing to physical discomfort.

The COVID-19 pandemic highlighted inadequate working conditions in many households due to ergonomic risk factors linked to lack of proper workstations, consistent with findings by El Kadri Filho & Roberto de Lucca (2022) (23). According to these authors, companies that maintain remote work should monitor working conditions and workload to prevent musculoskeletal disorders.

Regarding pain location, the most affected regions were the lumbar and cervical areas, corroborating findings from Argus & Pääsuke (2021) (14) and Santos et al. (2021) (20). These studies emphasize that sedentary and non-ergonomically adapted workers are particularly prone to pain in these areas. Moretti et al. (2020) also highlighted the widespread use of inadequate chairs and desks, contributing to pain, particularly spinal pain. Santos et al. (2021) (20) further reported a higher prevalence of cervical MSP among women in telework, possibly due to the dual burden of professional and domestic responsibilities.

Although most participants reported low coronaphobia, the results indicated a greater likelihood of pain with higher fear levels. This suggests a possible relationship between fear intensity and pain experience, reflecting the impact of emotional states on physical health. Celenay et al. (2020) (11) also reported higher pain and coronaphobia levels among home-office workers. Given that the present study’s sample was predominantly female, these findings align with Peres et al. (2021) (25) and Arpaci et al. (2021) (13), who reported greater female emotional vulnerability to COVID-19 threats. The significant correlation between fear and worsened MSP also suggests that fear influences not only mental health but also pain perception and processing, as proposed by Ahorsu et al. (2020) (26) and confirmed by Yıldırım et al. (2022) (27). This may indicate that ongoing stress and inadequate environments act synergistically, amplifying pandemic-related anxiety. Oakman et al. (2022) (28) found that poor workstation quality and high occupational demands were associated with more severe MSP, complementing the present findings.

This study also reinforces the importance of understanding coronaphobia as a multidimensional phenomenon, as highlighted by Vargová et al. (2023) (21), who differentiated between health-related and socioeconomic dimensions of pandemic anxiety.

Studies such as Oakman et al. (2022) (28) identified distinct MSP trajectories associated with factors such as workplace comfort, autonomy, and job demands. In this study, the association between work modality and MSP was statistically significant and consistent with existing literature. Moreover, Loef et al. (2022) (22) reported sedentary behavior as a mediator of the relationship between remote work and MSP, reinforcing the importance of interventions targeting sedentary lifestyles in home environments. Physical inactivity and sedentary behaviors increased across all Brazilian population groups during the pandemic (29), favoring the onset of pain conditions.

From a practical standpoint, the results highlight the need for institutional actions promoting regular physical activity during home-office, ergonomic adjustments to home workstations, and psychological support to reduce anxiety and coronaphobia.

Study limitations include its cross-sectional design, which precludes causal inference, and the use of a convenience sample, which limits generalizability. Future research should explore mediation between coronaphobia, coping strategies, and MSP in longitudinal models, as well as test interventions aimed at promoting mental and physical health in telework.

## CONCLUSION

This study revealed that during the COVID-19 pandemic, both physical and emotional health were significantly affected among Brazilian workers, particularly women and younger individuals.

In summary, changes in the work environment due to lockdown led to increased musculoskeletal pain. MSP prevalence rose after the pandemic onset, especially among individuals working from home. The most frequently reported pain sites were lumbar and cervical regions. Coronaphobia was associated with MSP, with individuals reporting moderate or high fear being more likely to experience pain.

The results underscore the importance of an integrated approach addressing physical, emotional, and environmental factors in promoting the health of home-office workers. The pandemic highlighted the urgent need to adapt work environments and provide emotional support to mitigate adverse effects on musculoskeletal health.

## Data Availability

All data produced in the present study are available upon reasonable request to the authors

## CONFLICT OF INTEREST

None to report.

## Notes

### Competing Interest Statement

The authors have declared no competing interest.

### Funding Statement

This study did not receive any funding

### Author Declarations

Research Ethics Committee of Universidade Paulista / UNIP gave ethical approval for this work (Protocol: 46136621.3.0000.5512).

